# Prolonged Dual Antiplatelet Therapy in Acute Myocardial Infarction Patients without Revascularization: Results from a China Acute Myocardial Infarction (CAMI) registry study

**DOI:** 10.1101/2023.08.22.23294450

**Authors:** Cunrong Huang, Jingang Yang, Ling Li, Shenghu He, Xuxia Zhang, Haiyan Xu, Yuan Wu, Jun Zhang, Shubin Qiao, Yongjian Wu, Yanyan Zhao, Yang Wang, Wei Li, Chen Jin, Xiaojin Gao, Yuejin Yang, CAMI (China Acute Myocardial Infarction) Registry Study Group

**Author notes:** C. Huang and J. Yang contributed equally. **Corresponding author** Xiaojin Gao, MD, and Yuejin Yang, MD; Complete address: Department of Cardiology, Coronary Heart Disease Center, Fuwai Hospital, Chinese Academyof Medical Sciences and Peking Union Medical College State Key Laboratory of Cardiovascular Disease, National Center for Cardiovascular Diseases,No. 167 N Lishi Rd, Xicheng District, Beijing, China. and.

## Abstract

**Background:** At least 12-month dual antiplatelet therapy (DAPT) is one of the standards of care following Percutaneous Coronary Intervention (PCI) in patients with acute coronary syndrome. However, study on prolonged DAPT for acute myocardial infarction (AMI) patients without revascularization is limited.

**Methods:** We studied 1744 AMI patients without revascularization from the China Acute Myocardial Infarction registry between January 2013 and September 2014. These patients were on DAPT and did not experience AMI, stroke, or bleeding events at 12-month follow-up. We divided them into two groups: 12-month DAPT group (DAPT for at least 12 months but less than 18 months) and 18-month DAPT group (DAPT for at least 18 months). The primary outcome is 24-month all-cause death.

**Results:** Overall, 1221 (70.0%) patients took DAPT for ≥12 months but <18 months, while 523 (30.0%) patients took DAPT for ≥18 months. The two groups had comparable proportions with high ischemic risk (27.0% vs. 25.6%, P = 0.5418), as well as high bleeding risk (29.0% vs. 28.5%, P = 0.8316). At 24 months, the all-cause mortality rate of 18-month DAPT group was significantly lower than that for 12-month DAPT group (3.7% vs 5.9%, P = 0.0471). Adjusted hazard ratio for all-cause death also showed statistical significance (0.59, 95% CI: 0.35-0.99, P = 0.0444).

**Conclusions:** DAPT for at least 18 months was associated with lower 24-month mortality for non-revascularization AMI patients without events within 12 months after onset.

---

**Registration:** URL: https://www.clini caltr ials.gov; Unique identifier: NCT01874691.

## Clinical Perspective

### What is new?

1. We provided the evidence that prolonging dual antiplatelet therapy (DAPT) to at least 18 months appears to reduce 24-month mortality risk in non-revascularization acute myocardial infarction patients without events within 12 months after onset.
2. We are concerned with myocardial infarction patients who were transitioning from an acute state to chronic stable coronary artery disease. This particular group of patients is large but often overlooked in most previous studies. Prolonged DAPT may provide them with additional clinical benefit.

### What are the clinical implications?

1. Extending DAPT for at least 18 months may be considered for non-revascularization myocardial infarction patients without events within 12 months after acute myocardial infarction onset.

### Non-standard Abbreviations and Acronyms

**DAPT** Dual AntiPlatelet Therapy

**CAMI** China Acute Myocardial Infarction

**PCI** Percutaneous Coronary Intervention

**AMI** Acute Myocardial Infarction

**MI** Myocardial Infarction

**ACS** Acute Coronary Syndrome

**STEMI** ST-segment Elevation Myocardial Infarction

**NSTEMI** Non ST-segment Elevation Myocardial Infarction

**MACCE** Main Adverse Cardiovascular and Cerebrovascular Event

## Introduction

Dual antiplatelet therapy (DAPT), the combination of aspirin and a P2Y12 inhibitor (clopidogrel, prasugrel, or ticagrelor), is a standard of care for the prevention of atherothrombotic events in all patients undergoing percutaneous coronary intervention (PCI), as well as those treated medically for acute coronary syndrome (ACS), but it also exposes patients to an increased bleeding risk.^1, 2^ Guidelines from Europe and the United States recommend that DAPT should be used at least 12 months for ACS patients undergoing PCI or managed with medical therapy.^3,4^ For patients with previous MI and are at low bleeding risk, latest guidelines and consensus suggested that extended DAPT beyond 12 months for a period of up to 3 years may be reasonable.^5,6^ However, most evidence mainly came from ACS patients who have undergone PCI, or the patients with a relatively long history (1∼3 years) of myocardial infarction (MI).^7,8^ Further study is still needed for acute myocardial infarction (AMI) patients who are treated without revascularization, as they may face a heightened risk of future plaque rupture relative to those without prior MI,^9^ indicating a potential need of prolonged DAPT for these patients. Using data from China Acute Myocardial Infarction (CAMI) registry, we investigated the association of prolonged DAPT and all-cause mortality in the AMI patients who did not experience any acute ischemic or bleeding events during the first year after MI and were still on DAPT at the 12-month follow-up.

## Methods

### Data Collection

Our study was conducted using data from the CAMI Registry between January 1, 2013 and September 30, 2014. The detail of the registry design was previously published.^10^ In brief, the CAMI Registry is a prospective, nationwide, multicenter observational study for AMI patients in China, collecting data on patients’ characteristics, treatments and prognoses. The CAMI Registry contained 108 hospitals at three levels (i.e., provincial, prefectural, and county levels), covering all the provinces and municipalities across mainland China, hence rendering it probably representative of routine real-world clinical setting of AMI care in China and reduce selection bias.^11^ This study was registered with ClinicalTrials.gov (NCT01874691) and approved by the institutional review board of all participating hospitals. Written informed consent was obtained from all patients enrolled or their relatives. The study protocol conforms to the ethical guidelines of the 1975 Declaration of Helsinki.^12^

### Study Population

Overall, 26,648 AMI patients from 108 hospitals were enrolled in the CAMI registry from January 1, 2013 to September 30, 2014. Exclusion criteria were as follows: (a) patients who could not be classified as “ST-segment elevation myocardial infarction (STEMI)” or “Non ST-segment elevation myocardial infarction (NSTEMI)”; (b) patients with primary or elective PCI and thrombolysis; (c) patients with main adverse cardiovascular and cerebrovascular events (MACCE) or bleeding events within 12 months; (d) taking DAPT < 12 months; Finally, 1744 AMI patients without revascularization were included in the present study (Figure 1).

**Figure 1.**
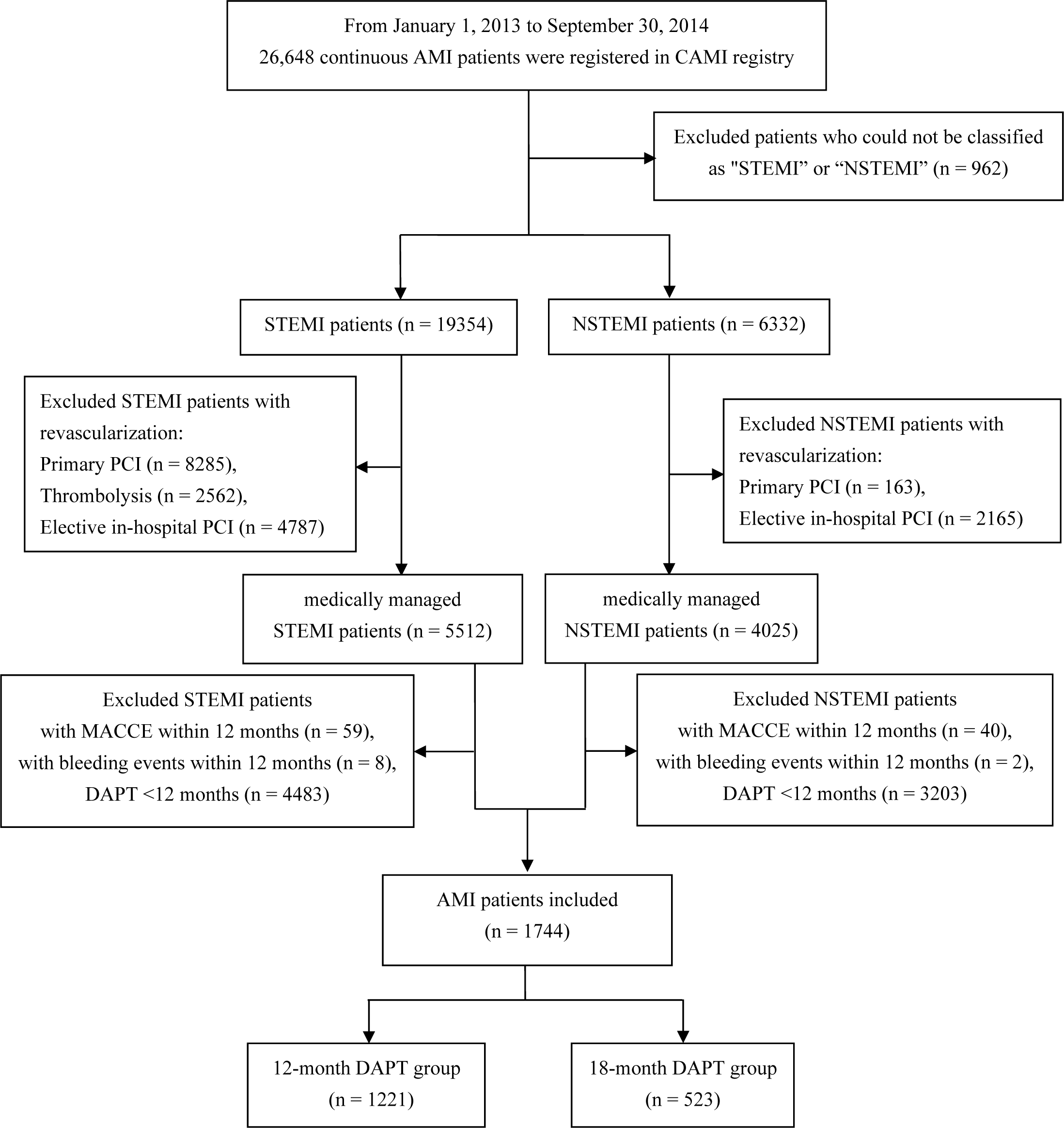
Flow chart for the included patients. AMI, acute myocardial infarction; CAMI, China Acute Myocardial Infarction; STEMI, ST-segment elevation myocardial infarction; NSTEMI, Non ST-segment elevation myocardial infarction; PCI, percutaneous coronary intervention; MACCE, main adverse cardiovascular and cerebrovascular event; DAPT: dual antiplatelet therapy

### Grouping and Definition

Diagnostic criteria of AMI conform to the third Universal Definition of Myocardial Infarction (2012).^13^ Patients who took DAPT at least 12 months but less than 18 months were assigned to 12-month DAPT group, and patients who took DAPT at least 18 months were assigned to 18-month DAPT group. The primary outcome was 24-month all-cause death (i.e., the death from any cause). In addition, we also analyzed some other outcomes, including MACCE which includes all-cause death, myocardial infarction (MI) or stroke, individual components of MACCE, and bleeding events (a composite of in-hospital cerebral hemorrhage, post cerebral infarction hemorrhage, and other hemorrhagic events) at both 18-month and 24-month follow-up.^11^ Standard definitions of the medical history and physical examination elements were well described in the ACC/AHA Task Force on Clinical Data Standards and the NCDR-ACTION-GWTG element dictionary.^14,15^ The Global Registry of Acute Coronary Events (GRACE) risk score before discharge was used to evaluate risk of ischemia for the participants, and higher score means higher ischemic risk. GRACE score >118 was defined as high ischemic risk.^16^ The CRUSADE (Can Rapid risk stratification of Unstable angina patients Suppress ADverse outcomes with Early implementation of the ACC/AHA Guidelines) score was used to evaluate bleeding risk for the patients, and higher score means higher bleeding risk. CRUSADE score ≥41 was defined as high bleeding risk.^17^ Subgroup analyses were based on prolonged DAPT (18 months≤DAPT<24 months or 24-month DAPT), age (<75 years or ≥75 years), diabetes (Yes or No), GRACE score (≤118 or >118), and CRUSADE score (<41 or ≥41). Follow-up data were obtained by local clinic visit, telephone interview, follow-up letter or public security system (for the participants who did not respond to the telephone interview or follow-up letter, public security system only helped to confirm whether the patient was alive or dead). All events were carefully checked and verified by an independent group of clinical physicians. Follow-up was scheduled at 1, 6, and 12 months, 18 months and 24 months after registered.

### Statistical Analysis

To evaluate the differences of baseline characteristics and clinical outcomes between the two treatment groups, continuous variables were presented as mean ± SD or median (25th–75th percentiles) and compared across groups by using t test. Categorical data was presented as counts and percentages and compared across groups by using Chi-square test or Fisher exact test and an analysis of variance. The differences in cumulative event rates were evaluated based on different durations of DAPT, using both Chi-square test and log-rank test, and its results were presented as Kaplan-Meier curves. Cox regression model was used to estimate hazard ratios (HRs) and 95% confidence interval (CI) for clinical outcomes mentioned above in relation to DAPT duration. Clinically or statistically significant covariates were entered into the multivariate Cox regression model. Missing adjusted covariates were imputed by maximum frequency for statistical analysis. There were 2 Cox models in total. Adjusted covariates for the Model_1_ included age, sex, diabetes, hypertension, type of MI, Killip class, anterior wall infarction, estimated glomerular filtration rate (eGFR), history of MI, and hospital level. For the Model_2_, besides the covariates of the Model_1_, we additionally included medications at discharge (statin, β Blocker, and angiotensin converting enzyme inhibitor/angiotensin II receptor blockers). For all analyses, a two-sided P-value of < 0.05 was considered significant. All statistical analyses were performed using SAS 9.4 (SAS Institute, Cary, NC, USA).

## Results

### Study Population

The details of participants’ baseline characteristics are shown in Table 1. Of 1744 AMI patients who did not undergo revascularization and were event-free within 12 months, 1221 (70.0%) patients had discontinued DAPT (12-month DAPT group) and 523 (30.0%) were still taking DAPT (18-month DAPT group) at 18 months. 12-month DAPT group had a higher proportion of patients aged≥75 years than 18-month DAPT group (23.4% vs 19.1%, P = 0.0451). The two groups had comparable proportions with high ischemic risk (27.0% vs. 25.6%, P = 0.5418), as well as high bleeding risk (29.0% vs. 28.5%, P = 0.8316). There were no significant differences in body mass index (BMI), hypertension, diabetes, type of MI, Killip class, estimated glomerular filtration rate (eGFR) between the two groups. Hospital level was unequally distributed between the two groups, with statistically significant differences (Provincial: 20.7% vs 33.7%, Municipal: 64.9% vs 55.6%, County: 14.3% vs 10.7%, P < 0.0001).

**Table 1.** Baseline clinical characteristics in all participants.

| **Characteristic** | **12-month DAPT group**<br>**(n = 1221)** | **18-month DAPT group**<br>**(n = 523)** | **P value** |
| --- | --- | --- | --- |
| **Age, yrs** | 64.5 ± 12.2 | 63.5 ± 12.3 | 0.1210 |
| **Age ≥ 75 (N, %)** | 286/1218 (23.5%) | 100/520 (19.2%) | 0.0451 |
| **Sex, Male (%)** | 858/1221 (70.3%) | 382/523 (73.0%) | 0.3116 |
| **BMI (kg/m<sup>2</sup>)** | 24.2 ± 3.2 | 24.1 ± 3.0 | 0.6487 |
| **Hypertension (N, %)** | 637/1200 (53.1%) | 296/519 (57.0%) | 0.1309 |
| **Diabetes (N, %)** | 249/1196 (20.8%) | 110/519 (21.2%) | 0.8608 |
| **Hyperlipidemia (N, %)** | 72/1198 (6.0%) | 26/519 (5.0%) | 0.4064 |
| **Current smoker (N, %)** | 450/1191 (37.8%) | 211/515 (41.0%) | 0.2156 |
| **Family CAD history (N, %)** | 39/1195 (3.3%) | 18/519 (3.5%) | 0.8288 |
| **History of angina (N, %)** | 404/1194 (33.8%) | 155/518 (29.9%) | 0.1112 |
| **History of MI (N, %)** | 121/1193 (10.1%) | 52/517 (10.1%) | 0.9576 |
| **History of heart failure (N, %)** | 48/1195 (4.0%) | 24/518 (4.6%) | 0.5626 |
| **History of Stroke (N, %)** | 110/1195 (9.2%) | 59/518 (11.4%) | 0.1687 |
| **History of PVD (N, %)** | 11/1194 (0.9%) | 8/517 (1.5%) | 0.2705 |
| **History of bleeding history (N, %)** | 24/1181 (2.0%) | 12/514 (2.3%) | 0.6939 |
| **History of liver disease (N, %)** | 9/1186 (0.8%) | 5/516 (1.0%) | 0.7708 |
| **History of chronic kidney (N, %)** | 15/1193 (1.3%) | 5/516 (1.0%) | 0.6045 |
| **History of PCI (N, %)** | 69/1194 (5.8%) | 40/516 (7.8%) | 0.1317 |
| **History of thrombolysis (N, %)** | 7/120 (5.8%) | 3/51 (5.9%) | 1.0000 |
| **History of CABG (N, %)** | 6/1194 (0.5%) | 7/518 (1.4%) | 0.0734 |
| **Type of MI (N, %)** |  |  | 0.3141 |
| **STEMI** | 681/1221 (55.8%) | 278/523 (53.2%) |  |
| **NSTEMI** | 540/1221 (44.2%) | 245/523 (46.8%) |  |
| **Anterior wall infarction (N, %)** | 531/1204 (44.1%) | 228/514 (44.4%) | 0.9224 |
| **Killip class ≥ II (N, %)** | 338/1194 (28.3%) | 144/517 (27.9%) | 0.8936 |
| **LVEF < 40% (N, %)** | 92/847 (10.9%) | 38/354 (10.7%) | 0.9483 |
| **Time from onset > 12 h (N, %)** | 588/1217 (48.3%) | 254/512 (49.6%) | 0.6609 |
| **WBC ($\times 10^9/L$)** | 9.5 ± 3.5 | 9.5 ± 3.3 | 0.9816 |
| **Hb < 120, g/L (N, %)** | 254/1165 (21.8%) | 109/506 (21.6%) | 0.9567 |
| **eGFR < 60, mL/min/1.73 m<sup>2</sup> (N, %)** | 374/1145 (32.7%) | 142/482 (29.5%) | 0.2265 |
| **LDL-C (mmol/L)** | 2.8 ± 1.1 | 2.8 ± 1.0 | 0.3289 |
| **GRACE Score before discharge (N, %)** |  |  | 0.4404 |
| ≤ 88 | 382/1221 (31.3%) | 180/523 (34.4%) | |
| 89-118 | 509/1221 (41.7%) | 209/523 (40.0%) |  |
| > 118 | 330/1221 (27.0%) | 134/523 (25.6%) |  |
| **High ischemic risk (Score>118)** | 330/1221 (27.0%) | 134/523 (25.6%) | 0.5418 |
| **CRUSADE Score (N, %)** |  |  | 0.7947 |
| ≤ 20 | 377/1221 (30.9%) | 164/523 (31.4%) | |
| 21-30 | 264/1221 (21.6%) | 106/523 (20.3%) |  |
| 31-40 | 226/1221 (18.5%) | 104/523 (19.9%) |  |
| 41-50 | 209/1221 (17.1%) | 81/523 (15.5%) |  |
| **> 50** | 145/1221 (11.9%) | 68/523 (13.0%) |  |
| **high bleeding risk (Score ≥ 41)** | 354/1221 (29.0%) | 149/523 (28.5%) | 0.8316 |
| **Medication (N, %)** |  |  |  |
| **Glycoprotein IIb/IIIa receptor blocker** | 186/1168 (15.9%) | 95/504 (18.8%) | 0.1456 |
| **Statin** | 1116/1133 (98.5%) | 470/483 (97.3%) | 0.1103 |
| **Nitrates** | 996/1198 (83.1%) | 415/512 (81.1%) | 0.3017 |
| **$\beta$ Blocker** | 823/1193 (69.0%) | 367/513 (71.5%) | 0.3014 |
| **Calcium channel blocker** | 214/1194 (17.9%) | 95/513 (18.5%) | 0.7699 |
| **ACEI/ARB** | 738/1192 (61.9%) | 313/513 (61.0%) | 0.7264 |
| **Hospital Level (N, %)** |  |  | <0.0001 |
| **Provincial** | 253/1221 (20.7%) | 176/523 (33.7%) |  |
| **Municipal** | 793/1221 (64.9%) | 291/523 (55.6%) |  |
| **County** | 175/1221 (14.3%) | 56/523 (10.7%) |  |

DAPT, dual antiplatelet therapy; BMI, body mass index; CAD, coronary artery disease; MI, myocardial infarction; PVD, peripheral vascular disease; PCI, percutaneous coronary intervention; CABG, coronary artery bypass grafting; STEMI, ST-segment elevation myocardial infarction; NSTEMI, Non ST-segment elevation myocardial infarction; LVEF, left ventricular ejection fraction; WBC, white blood count; Hb, hemoglobin; eGFR, estimated glomerular filtration rate; LDL-C, low-density lipoprotein cholesterol; ACEI, angiotensin converting enzyme inhibitor; ARB, angiotensin II receptor blockers

### Long-term Medications

At 12-month follow-up, there was no significant difference in adherence of secondary preventive medications between 12-month DAPT group and 18-month DAPT group. During the 18-month follow-up, there was a decrease in adherence to statins (86.4% vs 96.1%), β blockers (67.3% vs 71.4%), and ACEI/ARB (53.5% vs 57.5%) in the 12-month DAPT group compared with the 12-month follow-up. However, there was relatively minimal change in drug adherence observed in the 18-month DAPT group. At 24 months follow-up, only 194 (37.1%) patients in 18-month DAPT group still took DAPT. More information is demonstrated in Table 2.

**Table 2.** Long-term medications for 2 treatment groups.

|  | **12-month DAPT group** | **18-month DAPT group** | **P value** |
| --- | --- | --- | --- |
|  | **(n = 1221)** | **(n = 523)** |  |
| **Aspirin** |  |  |  |
| **12 months** | 1221/1221 (100.0%) | 523/523 (100.0%) | - |
| **18 months** | 738/805 (91.7%) | 523/523 (100.0%) | <0.0001 |
| **24 months** | 635/692 (91.8%) | 374/400 (93.5%) | 0.2916 |
| **P2Y12 inhibitor** |  |  |  |
| **12 months** | 1221/1221 (100.0%) | 523/523 (100.0%) | - |
| **18 months** | 12/804 (1.5%) | 523/523 (100.0%) | <0.0001 |
| **24 months** | 35/690 (5.1%) | 198/399 (49.6%) | <0.0001 |
| **DAPT** |  |  |  |
| **12 months** | 1221/1221 (100.0%) | 523/523 (100.0%) | - |
| **18 months** | 0/1221 (0%) | 523/523 (100.0%) | <0.0001 |
| **24 months** | 0/1221 (0%) | 194/523 (37.1%) | <0.0001 |
| **Statin** |  |  |  |
| **12 months** | 1172/1220 (96.1%) | 504/522 (96.6%) | 0.6234 |
| **18 months** | 696/806 (86.4%) | 503/521 (96.5%) | <0.0001 |
| **24 months** | 587/692 (84.8%) | 364/400 (91.0%) | 0.0027 |
| **β Blocker** |  |  |  |
| **12 months** | 867/1214 (71.4%) | 370/518 (71.4%) | 0.9960 |
| **18 months** | 541/804 (67.3%) | 367/517 (71.0%) | 0.1560 |
| **24 months** | 459/690 (66.5%) | 265/396 (66.9%) | 0.8936 |
| **ACEI/ARB** |  |  |  |
| **12 months** | 695/1208 (57.5%) | 283/509 (55.6%) | 0.4601 |
| **18 months** | 431/805 (53.5%) | 278/511 (54.4%) | 0.7596 |
| **24 months** | 358/690 (51.9%) | 187/392 (47.7%) | 0.1862 |

DAPT, dual antiplatelet therapy; ACEI, angiotensin converting enzyme inhibitor; ARB, angiotensin II receptor blockers

### Clinical Outcomes

At 18-month follow-up, 18-month DAPT group had significantly lower rate of all-cause death (1.1% vs 3.4%, P = 0.0048) and MACCE (1.9% vs 4.3%, P = 0.0096) than 12-month DAPT group (Table 3). In the Cox regression analysis, adjustments were made for statistically or clinically significant covariates (Table 4), and it was found that the 18-month DAPT was significantly associated with a reduction in the all-cause death risk, regardless of whether covariates regarding medications were included in the Cox model (adjusted hazard ratio_2_ [aHR], 0.32 (95% CI, 0.13-0.76), P = 0.0097).

**Table 3.** Cumulative event rates for 2 treatment groups at 18-month and 24-month.

| **Outcomes** | **12-month DAPT group**<br>**(n = 1221)** | **18-month DAPT group**<br>**(n = 523)** | **P value** |
| --- | --- | --- | --- |
| **18-month follow-up** |  |  |  |
| **All-cause death** | 40/1188 (3.4%) | 6/523 (1.1%) | 0.0048 |
| **MACCE** | 51/1188 (4.3%) | 10/523 (1.9%) | 0.0096 |
| **MI** | 7/1164 (0.6%) | 4/523 (0.8%) | 0.7467 |
| **Stroke** | 5/1163 (0.4%) | 0/522 (0.0%) | 0.3322 |
| **Bleeding events** | 1/1163 (0.1%) | 0/523 (0.0%) | 1.0000 |
| **24-month follow-up** |  |  |  |
| **All-cause death** | 70/1179 (5.9%) | 19/517 (3.7%) | 0.0471 |
| **MACCE** | 88/1179 (7.5%) | 28/517 (5.4%) | 0.1164 |
| **MI** | 9/1158 (0.8%) | 8/511 (1.6%) | 0.1541 |
| **Stroke** | 10/1157 (0.9%) | 2/511 (0.4%) | 0.3643 |
| **Bleeding events** | 3/1157 (0.3%) | 1/511 (0.2%) | 1.0000 |

DAPT, dual antiplatelet therapy; MACCE, main adverse cardiovascular and cerebrovascular event; MI, myocardial infarction

**Table 4.** Hazard ratios of different follow-up time for all-cause death and MACCE (the reference group is 12-month DAPT group)

| **Outcome** | **aHR<sub>1</sub><sup>\*</sup> (95% CI)** | **P<sub>1</sub> value** | **aHR<sub>2</sub><sup>†</sup> (95% CI)** | **P<sub>2</sub> value** |
| --- | --- | --- | --- | --- |
| **All-cause death** |  |  |  |  |
| **18-month all-cause death** | 0.33 (0.14-0.79) | 0.0124 | 0.32 (0.13-0.76) | 0.0097 |
| **24-month all-cause death** | 0.60 (0.36-1.00) | 0.0517 | 0.59 (0.35-0.99) | 0.0444 |
| **MACCE** |  |  |  |  |
| **18-month MACCE** | 0.47 (0.24-0.94) | 0.0315 | 0.47 (0.23-0.93) | 0.0297 |
| **24-month MACCE** | 0.74 (0.48-1.14) | 0.1727 | 0.74 (0.48-1.14) | 0.1692 |

aHR, adjusted hazard ratio; CI, confidence interval; MACCE, main adverse cardiovascular and cerebrovascular event;
\* Adjusted covariates for aHR<sub>1</sub> included age, sex, diabetes, hypertension, type of myocardial infarction (MI), Killip class, anterior wall infarction, estimated glomerular filtration rate (eGFR), history of MI, and hospital level.
† Adjusted covariates for aHR<sub>2</sub> included age, sex, diabetes, hypertension, type of myocardial infarction (MI), Killip class, anterior wall infarction, estimated glomerular filtration rate (eGFR), history of MI, medication at discharge (statin, $\beta$ Blocker, and angiotensin converting enzyme inhibitor/angiotensin II receptor blockers), and hospital level.

At the 24-month follow-up, the patients that received ≥ 18 months DAPT still demonstrated a significantly lower all-cause death rate (3.7% vs 5.9%, P = 0.0471) compared to the group that received DAPT ≥12 months but <18 months (Table 3). There was no significant evidence of the continuation of DAPT (≥ 18 months) with respect to a higher rate of bleeding events relative to 12-month DAPT group (0.2% vs 0.3%, P = 1.0000). In addition, in the multivariable Cox regression analysis, prolonged DAPT (18-month DAPT group) was also significantly associated with a decreased risk of all-cause death at 24-month follow-up [aHR_2_, 0.59 (95% CI, 0.35-0.99), P = 0.0444]. Kaplan-Meier curves for all-cause death and MACCE stratified by the DAPT duration were shown in Figure 2.

**Figure 2.**
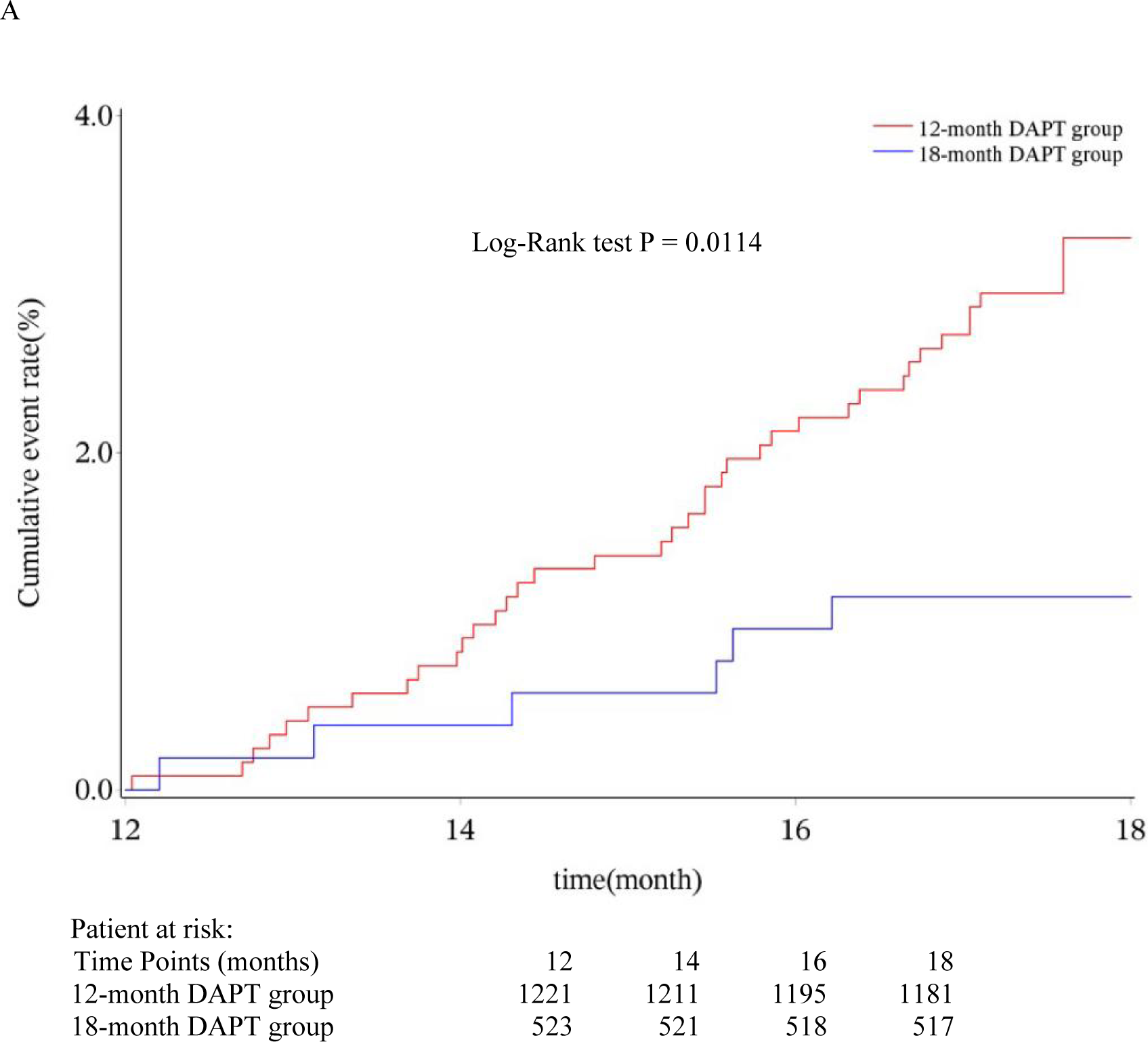
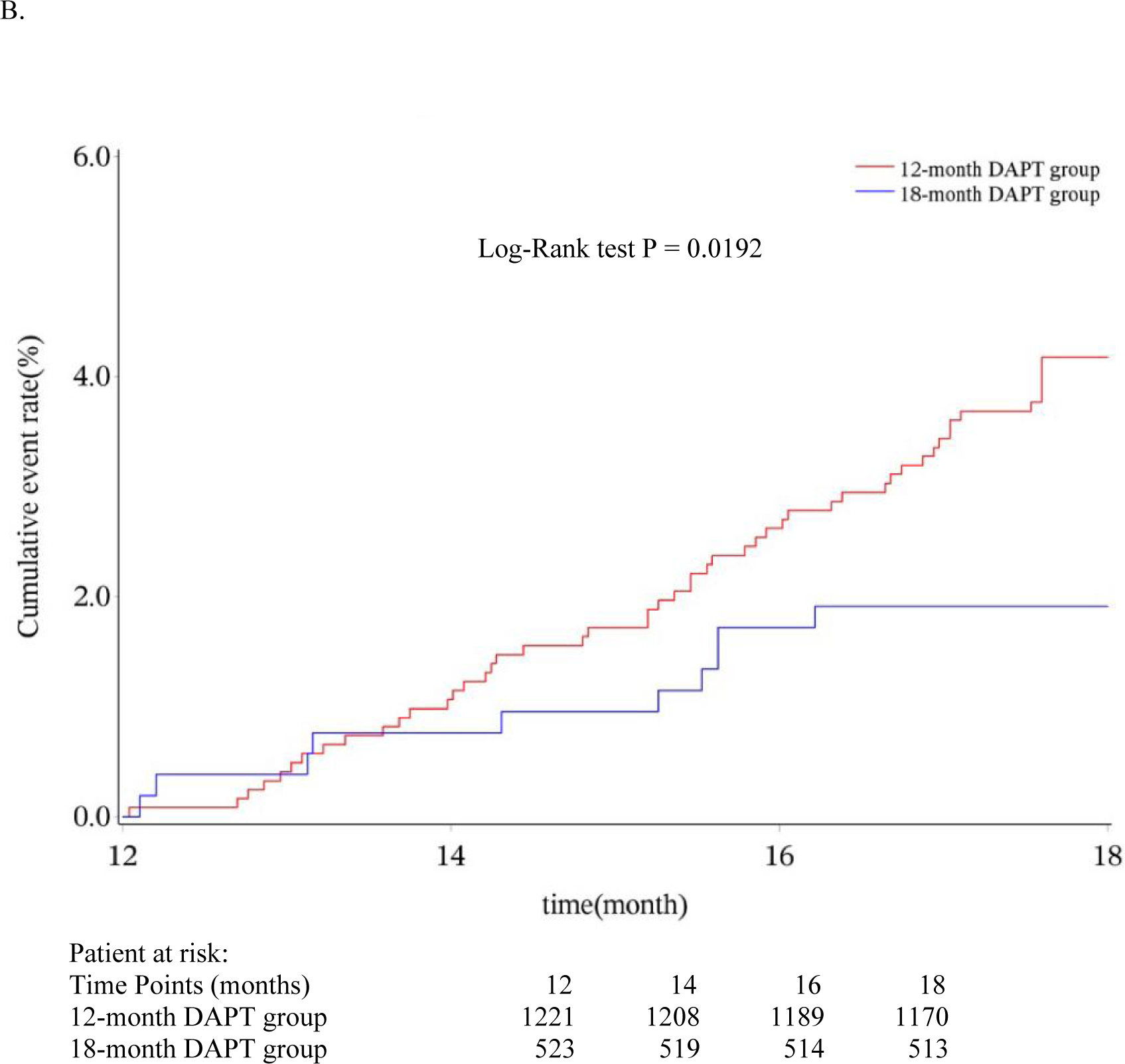
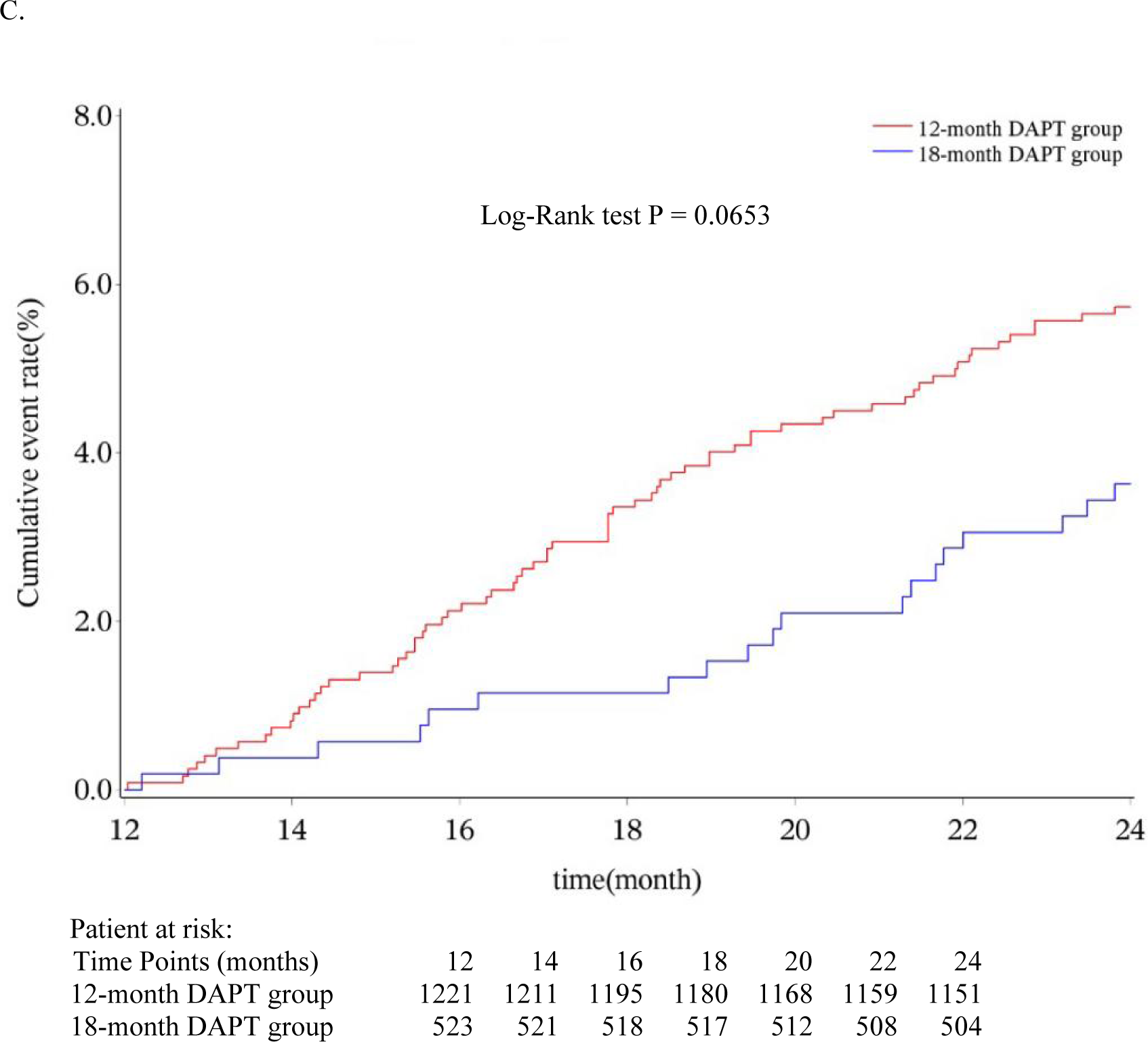
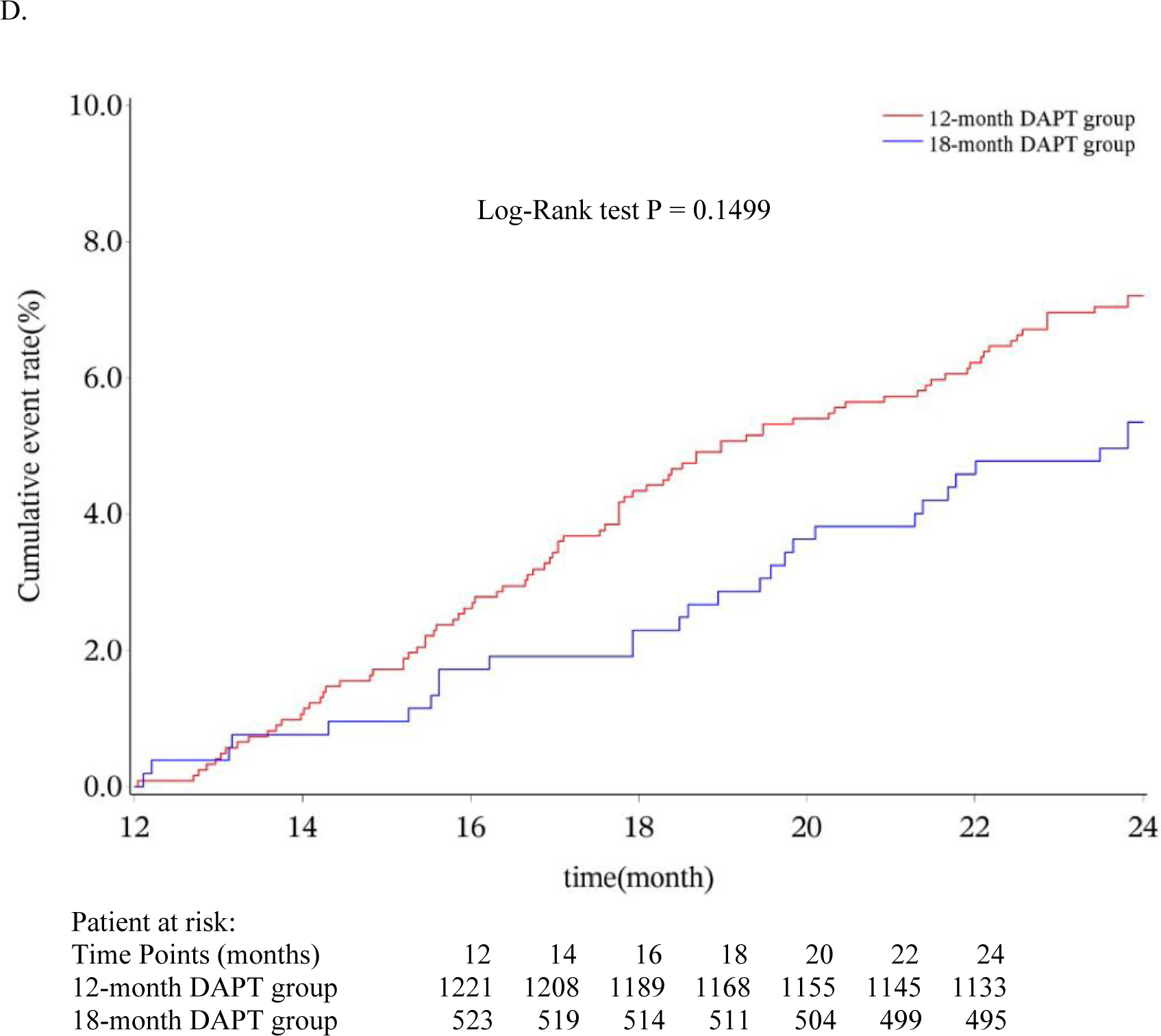
Kaplan-Meier curves and log-rank test for all-cause death and MACCE stratified by the DAPT duration. A.18-month all-cause death. B. 18-month MACCE. C.24-month all-cause death. D. 24-month MACCE. DAPT, dual antiplatelet therapy; MACCE, major adverse cardiovascular and cerebrovascular event.ss

### Subgroup analysis

Results of subgroup analysis based on prolonged DAPT were shown in Table S1. Patients on continuous DAPT (24 months) had significantly lower cumulative event rates of all-cause death and MACCE at 24 months compared with patients on 18 months ≤ DAPT < 24 months (all-cause death: 0.5% vs 5.6%, P = 0.0007; MACCE: 2.6% vs 7.1%, P = 0.0198). Kaplan-Meier curves for 24-month all-cause death and MACCE in the subgroups were presented in Figure S1. Log-rank test results also indicated that cumulative rate of both all-cause death (P = 0.0035) and MACCE (P = 0.0315) were significantly lower in 24-month DAPT subgroup than in 18 months ≤ DAPT < 24 months subgroup.

In addition, subgroup analysis also showed that there were no significant interactions between DAPT duration and age, diabetes, GRACE score, and CRUSADE score for 24-month all-cause death. However, among the participants aged <75 years and the non-diabetic patients, 18-month DAPT group seemly reduced the risk of 24-month all-cause death relative to 12-month DAPT group. More details were demonstrated in Figure S2.

## Discussion

In this multicenter, prospective AMI patient registry, we included patients who did not receive revascularization and were event-free (events included death, MI, stroke, or bleeding event) within 1 year from the symptom onset. In our study, prolonged DAPT (18-month DAPT group) had a significantly lower cumulative all-cause death rate than 12-month DAPT group at both 18 months and 24 months. Compared with the long-term DAPT (12 months), there was no significant increase in bleeding risk for the prolonged DAPT. Our study of MI patients without revascularization fills a research gap on whether there is any additional benefit to continuing DAPT after taking DAPT for a full 12 months in such patients, and concluded that continuing DAPT for at least 18 months was probably associated with a reduction in the risk of all-cause death compared with discontinuation of DAPT.

First, the participants we included were unique since they were the patients who actually transitioned from an acute state to chronic stable coronary artery disease (CAD), although they did not strictly meet the general definition of “stable post-MI populations” (i.e., the time between patients’ last MI or coronary intervention and inclusion was more than 1 year). PEGASUS-TIMI 54 trial,^18^ a landmark double-blind randomized controlled trial (RCT) on patients aged ≥ 50 years who had experienced a MI between 14.4 and 27.6 months ago, demonstrated that the rate of composite endpoints of cardiovascular death, MI, or stroke in DAPT group was around 3.5%, while the rate of composite endpoints was 4.5% in placebo+aspirin group at 18-months. The subgroup analysis showed long-term DAPT reduced thrombotic events in patients with prior MI, regardless of whether they had prior coronary stenting.^19^ Similar to PEGASUS-TIMI 54 trial, the MACCE rate in our study was 1.9% in 18-month DAPT group and 4.3% in 12-month DAPT group, supporting that the continuous DAPT may reduce the risk of ischemic events. Additionally, CHARISMA trial (40.6% participants of the subgroup had prior MI) showed that the rate of composite outcomes of cardiovascular death, MI, or stroke at 18 months was 4.5% in DAPT group and 6% in aspirin+placebo group, and appeared to derive a benefit of 17% deduction in the risk of cardiovascular death, MI, or stroke from DAPT with clopidogrel plus aspirin during the 28-month follow-up ^20^ Overall, the previous published studies on stable CAD directly included the patients with a history of MI, and their MI histories span a wide range of time, from over 1 year to around 30 years, and treatments patients taken were different, such as PCI, coronary-artery-bypass-grafting (CABG) or medical management. Thus, in fact, their populations were very heterogeneous which means the overall effect derived from the studies is a relatively rough average, overestimating the effect for some patients and underestimating the effect for another participants. On the contrary, our study was a continuation of the patients with AMI (exactly 1 year ago), and in our population, patients who underwent primary, elective interventions, and thrombolysis were all excluded, which means our subjects were a determined MI population, with the use of medication exclusively. Therefore, our findings are relatively more precise due to the clarity of our target population.

Second, we used all-cause death rather than cardiovascular death as the endpoint because our death records were mostly from death registration or public security system. In this real-world observational cohort, all-cause death record has relatively higher accuracy and data completeness. There were some other studies of DAPT using all-cause death as their study outcome as well.^21,22^ Thus, in relation to previous studies, our study mainly focused on all-cause death and drew a conclusion for this “hard endpoint” in MI patients. Comparing our results to the studies where the study populations was stable post-MI patients, CORONO study conducted by Delsart P et al. showed that the rate of all-cause death for their stable CAD patients without coronary vascular intervention history was around 5.5% at 1 year,^23^ which was higher than the corresponding rate in our 18-month DAPT group (3.7%) and similar to the rate in our 12-month DAPT group (5.9%). The baseline rate of DAPT use for their non-coronary vascular intervention patients was 20%, and angiotensin system antagonists, beta-blockers and statins were widely prescribed in these patients. According to the results, DAPT continued for at least 18 months is likely to reduce all-cause death rate for stable MI patients without revascularization because our patients had a shorter period of time since their last MI onset and should have been at a relatively higher risk of death compared to the stable patients with a history of MI >1 year,^9,24^ but as far as the current results are concerned, our patients who consistently used DAPT (18-month DAPT group) had a lower all-cause death rate, indicating a potential benefit of prolonged DAPT.

Third, in terms of bleeding risk for prolonged DAPT, unlike the results of other studies which found that prolonged DAPT was associated with increased rate of major bleeding events, our study showed that prolonged DAPT (18 months) did not significantly increase the risk of bleeding relative to long-term DAPT (12 months) in our participants. This inconsistency may be related to the following reasons. (1) It is known that several factors such as advanced age (> 65 years), low body weight, diabetes, or chronic kidney disease may increase bleeding risk.^25^ However, as shown in our baseline analysis, only a minority of participants in our study had risk factors noted above and these variables were distributed without significant difference between our two treatment groups, which may account for the inconsistent findings regarding bleeding risk. (2) We have excluded the patients who had major bleeding events within 12 months when we included the participants, which also reduced the bleeding risk in our study population to some extent. In fact, the incidence of bleeding events was very low in both of our 2 treatment groups, with 3 (0.3%) and 1 (0.2%) case for long-term DAPT group and prolonged DAPT group, respectively, at 24 months. Therefore, prolonged DAPT may not significantly increase bleeding events in the MI patients with a relatively low bleeding risk. A study carried out by Costa F et al. found that the bleeding risk was increased by long-term DAPT (12 or 24 months) only in the high-bleeding-risk patients, which also provided support to our results.^26^

Finally, in our study, although there was still a significant reduction in all-cause death risk for 18-month DAPT group relative to 12-month DAPT group at 24 months, the corresponding adjusted HR was slightly higher than the adjusted HR at 18 months, indicating a slight decline in the effect of protection. It may be related to the following reason. At 18 months follow-up, DAPT use was significantly better in 18-month DAPT group than in 12-month DAPT group (i.e., the rate of aspirin and P2Y12 inhibitor usage was both 100% for patients in 18-month DAPT group, while the rate of P2Y12 inhibitor usage in 12-month DAPT group was only 1.5%). However, at 24 months, the actual use of DAPT was only 194 persons (37.1%) in 18-month DAPT group. Thus, in fact, the majority of patients in 18-month DAPT group strictly used DAPT for only 18 months, and this result indicated that discontinuation of DAPT seemly means a reduction in cardiovascular protection to some extent. Moreover, for further exploring potential benefit of 24-month DAPT, we carried out the subgroup analysis (shown in Table S1) and found that the 24-month cumulative event rate of all-cause death did occur significantly less frequently in the subgroup of 24-month DAPT (i.e., strictly continuing DAPT at least 24 months) than in the subgroup of 18 months ≤ DAPT < 24 months (i.e., probably discontinuing after 18 months), which complemented and supported the additional benefit of continuous DAPT. Moreover, compared with other published studies, the 95% CI of our results of HR was relatively wider, possibly because the number of MI patients without revascularization in clinical study is very limited, and in developing countries, such as China, even fewer patients follow medical advice and are on long-term DAPT (at least 12 months), resulting in our relatively small sample size. Small sample size may lead to the instability of effect to some extent, presenting as a wide CI. However, it is important to note that even with our small sample size, we still got a statistically significant result, suggesting that there is indeed some protection effect from prolonged DAPT. In the future, the ideal sample size required to test effects of the treatment may be met by including additional high-risk populations.^27^

### Limitations

There are some limitations in our study. First, almost all of the P2Y12 inhibitors used in our study were clopidogrel, thus the results may not reflect influence of other P2Y12 inhibitor drugs. Second, the cause of death could not be determined as it was mostly collected from death registration. This is one of the reasons we did not select MACCE as our primary study outcome as well. Also, our registry study included hundreds of hospitals at different levels, making the collection of events other than death relatively less complete. Third, although we have adjusted several potential confounders between the two treatment groups, including medication, the actual medication use of the patients would not remain exactly constant during the follow-up period, which is a common limitation of observational studies, and it is also likely that there are several other unknown factors that may contribute to selection bias. Fourth, the records of important post-discharge events were mainly from the descriptions of patients and their relatives, which may cause recall bias. In addition, a large number of patients in China did not use DAPT to 1 year, not only leading to small sample size of our study, but also probably resulting in the occurrence tendency test unreliable (with a relatively wide confidence interval).

## Conclusion

Medically managed patients with AMI were not included in most current clinical trials, and therefore evidence for the treatment of the category of the patients are lacking. DAPT for at least 18 months was associated with lower mortality for non-revascularization AMI patients without events within 12 months after onset relative to 12-month DAPT. A more definitive impact of prolonged DAPT on long-term prognosis needs to be validated in the future with larger cohort studies. Notably, it is necessary to assess the patient’s bleeding risk before using prolonged DAPT strategy.

In PCI era, although MI patients without revascularization are less probable to receive evidence-based medications, this actually occurs more often when they are cared for by non-cardiologists, and it is one of reasons they became the target population of our study. ^28,29^ There have been some DAPT-related studies focusing on MI patients without revascularization. For instance, the CURE trial conducted by Yusuf S et al, a study that more than 60% of its patients with ACS did not undergo coronary revascularization, showed that 3∼12 months DAPT led to a 20% relative risk reduction of the composite endpoints of cardiovascular death, MI or ischemic stroke, which suggested the necessity of DAPT for medically managed patients,^30^ as in our study. Moreover, another large retrospective cohort study also revealed the effectiveness of DAPT for this kind of population.^22^ The evidences thus far reveal that taking DAPT at least 12 months has a significant mortality deduction and a protective effect against ischemic complications in MI patients, regardless of the presence of ST elevation and whether reperfusion is undergone or not.^31^ Under this situation, a key strength of our real-world cohort is its ability to examine our particular populations and to generalize our new findings of prolonged DAPT to real-world patients and practice settings.

## Data Availability

All data referred to in the manuscript is available.

## Acknowledgments

Jingang Yang and Cunrong Huang developed the idea and designed the study; Shenghu He, Xuxia Zhang, Haiyan Xu, Yuan Wu, Jun Zhang, Yongjian Wu, and Chen Jin collected the data; Ling Li, Yanyan Zhao, Yang Wang, and Wei Li carried out data analysis; Cunrong Huang wrote the first manuscript and all the authors have reviewed it; Xiaojin Gao, Shubin Qiao, and Yuejin Yang are the guarantors. All the authors claim that listed authors meet the authorship criteria and no one who meets the criteria is left out.

## Supplemental Material

Table S1 Figures S1-S2

